# Latiné immigrant heterogeneity: Striking health differences among Cuban refugee/migration waves to the United States

**DOI:** 10.1101/2024.04.17.24305856

**Authors:** Margarita Hernandez, George H. Perry

## Abstract

Latiné people differ markedly in our lived experiences in ways that are underappreciated. Meanwhile, variations in social experiences are known to be associated with differential health outcomes. We test whether immigration history is associated with health differences among U.S.-based Cuban refugees. Cubans from the circum-1980 Mariel Boatlift migration wave reported significantly higher instance and severity of disability on average than Early Cuban Exiles (1959-1962), Freedom Flight refugees (1965-1973), and Special Period (1990-2002) refugees. In contrast, disability among immigrants from Mexico to the U.S. across time-matched periods followed a different pattern. To help identify potential factors underlying the Cuban refugee pattern, we interviewed Miami-based Cubans. Participants described heightened discrimination in 1980s Cuba and U.S., which we hypothesize contributed to higher instances of disability refugees of that era. Our results suggest that understanding the history of Latiné immigration relative to country of origin, and the differential experiences of individuals both across and among ethnic groups, is imperative. A more nuanced understanding of the social determinants of immigrant health is important to help combat adverse factors in future generations.

**Significance Statement:** Even within a Latiné community from a single country of origin, we find that differential social experiences of immigrating to the U.S. are associated with variation in health outcomes. Understanding the social determinants of health within marginalized populations can help scientists, policymakers, and community-based organizers contextualize the conditions in which adverse health outcomes are arising and organize to combat these factors in future generations of refugees and immigrants coming to the United States.

## Introduction

Severe health disparities exist among individuals of different racial groups, ethnicities, sexes, genders, sexual orientations, and socioeconomic statuses in the United States (*1, 2*). Limited access to health resources (*3*), experienced marginalization (*4*), discrimination (*5*), and various other socioeconomic factors are associated with higher rates of many diseases in minority populations (*6*). The social conditions under which health manifests are commonly referred to as the social determinants of health (*7*).

Racial and ethnic groups are highly heterogeneous in their social experiences. Yet these groups are often homogenized in both public discourse and scientific literature, undermining recognition of the potential contributions of this heterogeneity to morbidity and mortality. For example, terms like Hispanic and/or Latiné refer to a broad group of people who trace their ancestry to Spanish-speaking Latin-American countries and are composed of individuals from different genetic, socioeconomic, and sociopolitical backgrounds (*8*). Yet within the United States, Latiné populations have traditionally been treated and studied as a monolithic group (*9*).

In this study, we assessed how immigration history and lived experiences factor into health outcomes among Cuban refugees in the United States. We asked: 1) Do health outcomes vary within and among different migration waves of Cubans to the United States? and 2) Are differential health outcomes within migration waves due to different lived experiences and environmental contexts? By understanding how differences in social experiences may relate to differences in health within one refugee/immigrant group, we modeled the importance of investigating heterogeneity within minority populations in the United States to provide a rich and nuanced understanding of the social determinants of health and the ways these adverse experiences can be combated.

### Cuban migration to the United States

The movement of Cubans from Cuba to the United States has a long history, often punctuated by political events and economic conditions in Cuba (*10*). Although there was Cuban migration to the United States prior to the Cuban Communist Revolution, totaling nearly 180,000 immigrants between 1896 and 1950, that migration rate was relatively dwarfed by that once the Communist regime took over the Cuban government (*11*). Post-revolutionary Cuban migration is often described as occurring in waves, each comprising the movements of individuals of distinct occupations, political positions, socioeconomic statuses, and races of Cubans to the United States (*12*–*15*).

The waves of interest for this study include: 1) Early Cuban Exiles (1959-1962), 2) Freedom Flights (1965-1973), 3) Mariel Boatlift (1980), and 4) The Special Period (1990 through the early 2000s). As Pedraza-Bailey (*15*) notes, “overall, the Cuban migration is characterized by an inverse correlation between date of departure and social class of the immigrants.” Even more severely, Bach (*16*) writes, “…there has been a total transplantation of the pre-revolutionary Cuban social structure to Miami, with all the implications of unequal wealth, power, and prestige.”

The Early Cuban Exiles were the first group of Cubans to leave after the Cuban Communist Revolution in 1959. These individuals were predominantly White, upper-, upper-middle, and middle-class Cubans who were the first beneficiaries of the Cuban Refugee Program, which, in part, sought to retrain medical professionals, teachers, and lawyers to be able to successfully continue their occupations within the United States (*13, 17*).

The second wave of migration, the Freedom Flights, occurred between 1965 and 1973, during which time Cubans with family members who had immigrated to the United States could leave the country and reunite with their relatives abroad (*13*). This immigration wave was primarily composed of individuals who were predominantly White and working-class.

In 1980, the Cuban government opened the port of Mariel in response to the Peruvian Havana Embassy Crisis of 1980, which allowed exiled Cubans to return and pick up their family members (*12*). Additionally, the Cuban government expelled individuals they deemed as “scum”, including any imprisoned individuals, those within psychiatric facilities, sex workers, and those who self-identified as queer (*13, 14*). Cubans who came to the United States during the Mariel Boatlift were predominantly working-class and were more racially and ethnically diverse than previous waves, with this wave containing the highest percentage of Black and *mestizo* Cuban immigrants (*19*).

The last wave - the Special Period - was disproportionately composed of Cubans of low-socioeconomic status (*20, 21*). This wave did not include proportionally as many Afro-Cubans and *mestizos* as the Mariel Boatlift wave, and individuals typically traveled by rafts across the Caribbean Sea from Cuba to the Florida Keys (*12*).

## Results

We aimed to test whether social experience heterogeneity is associated with differential health outcomes experienced by Cuban refugees living in the United States. To do so, we analyzed survey data from Cuban American participants in The National Latino and Asian American Study (NLAAS). The NLAAS is a population-based survey of Latiné and Asian American individuals across the United States, including Alaska and Hawai’i (*23*). The purpose of the NLAAS is to collect information on the mental health needs and services used by individuals of Latiné and Asian descent, with a variety of additional data collected, including on socioeconomic variables, experiences of discrimination, and on physical health (*24*).

Additionally, we applied the same analytical methodology (including with the same immigration period years, even though the social periods relevant for Cuban immigration are not necessarily relevant for other immigrant populations) with Mexican participant data within the NLAAS to test whether patterns observed among Cuban immigration waves are more likely explained by factors specific to Cubans or by general U.S.-wide factors that would have been experienced by all Latiné people.

### Cuban Participants within NLAAS

577 individuals within the NLAAS dataset at the time of our analysis were categorized as Cuban. We removed individuals from the sample that were born in the United States (N=76) and those whose migration year was ambiguous for wave categorization (N=25). We categorized individuals based on the time frame when they arrived in the United States, with 87 individuals categorized as Early Cuban Exiles (1953-1964), 105 individuals categorized as Freedom Flights (1965-1975), 106 individuals categorized as Pre- and Post-Mariel Boatlift (1977-1987), and 168 individuals categorized as the Special Period (1991-2002) (Figure 1).

**Figure 1.**
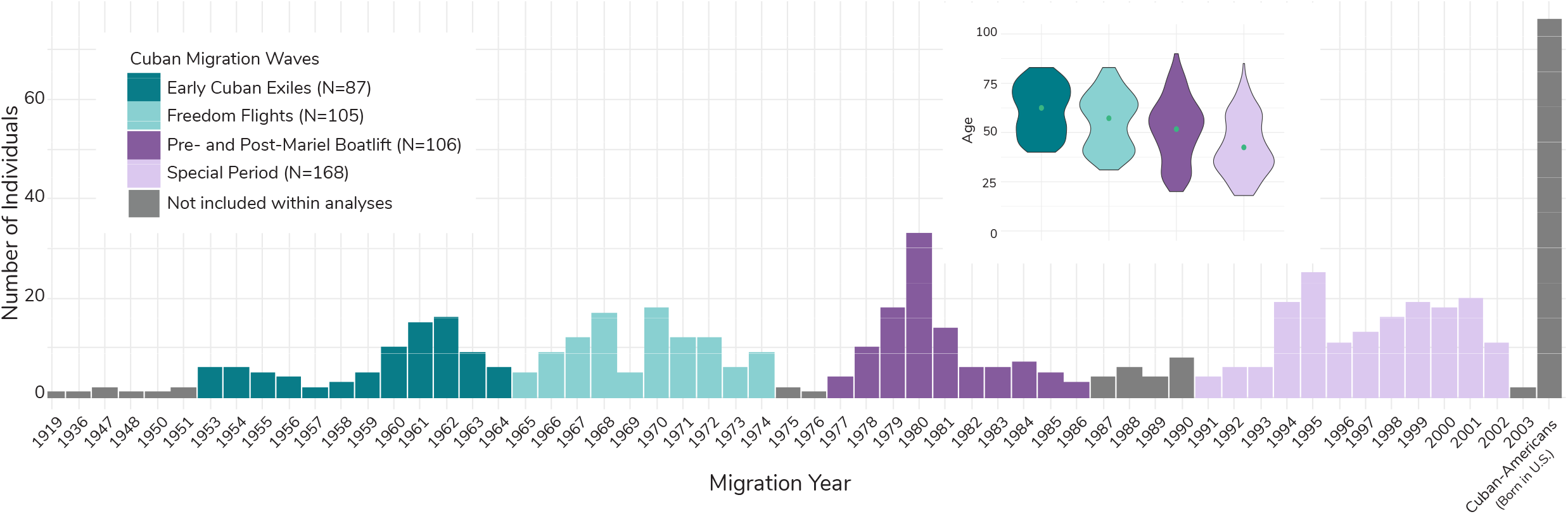
Number and average age of individuals of Cuban descent and their migration years (if applicable) within NLAAS. A) This chart shows the number of individuals of Cuban descent that came to the United States between 1919 and 2003. The colors correspond to the migration waves of interest for this study, with dark teal representing the Early Cuban Exiles, light teal representing the Freedom Flights, dark purple representing Pre- and Post-Mariel Boatlift, and light purple representing the Special Period. Individuals represented in gray were not included in downstream analyses, as they did not fit within the confines of the migration waves of interest for the study. The right-most bar represents the number of first- and second-generation Cuban-Americans within the study, which were also excluded from further analyses. Note: Within this study, Cuban migration to the United States in the 1910s through 1950 was very low. The years in which there are no migrants coming from Cuba to the United States are omitted from the chart. However, migration became consistent after 1953. The inset contains violin plots of mean age (at the time of data collection) and distribution for each Cuban refugee/migration wave as part of this study. Green dots represent the weighted mean for each wave, calculated using NLAAS sampling weights.

### Pre- and Post-Mariel Boatlift immigrants are more disabled on average than those from other waves

There were marginally significant differences in average self-care, cognition, mobility, and social interaction scores across Cuban refugee/migration waves, with individuals from the Pre-and Post-Mariel Boatlift having consistently higher scores across all domains (Supplementary Figure 2). The Pre- and Post-Mariel wave also had significantly and >2x higher role functioning disability scores than the other migration waves. Health score averages and standard deviations across migration waves are reported in Supplementary Figure 2.

Individuals from the Pre- and Post-Mariel Boatlift wave also had sum disability scores (41.59 ± 81.76) that were 2.4x higher than the Early Cuban Exiles (17.14 ± 45.82, ANCOVA; *p*=0.020), 2x higher than the Freedom Flights (20.58 ± 53.38, *p*=0.045), and 2.5x higher than the Special Period (16.41 ± 44.13, *p*=0.0030) (Figure 2, Supplementary Figure 2). We also plotted linear models for sum disability score against age for each migration wave (Figure 2B) and noted the stark upward shift of the Pre- and Post-Mariel Boatlift model relative to the other migration waves, with the youngest individuals within that wave having similar disability scores as the oldest members of the Early Cuban Exiles and the Freedom Flights.

**Figure 2.**
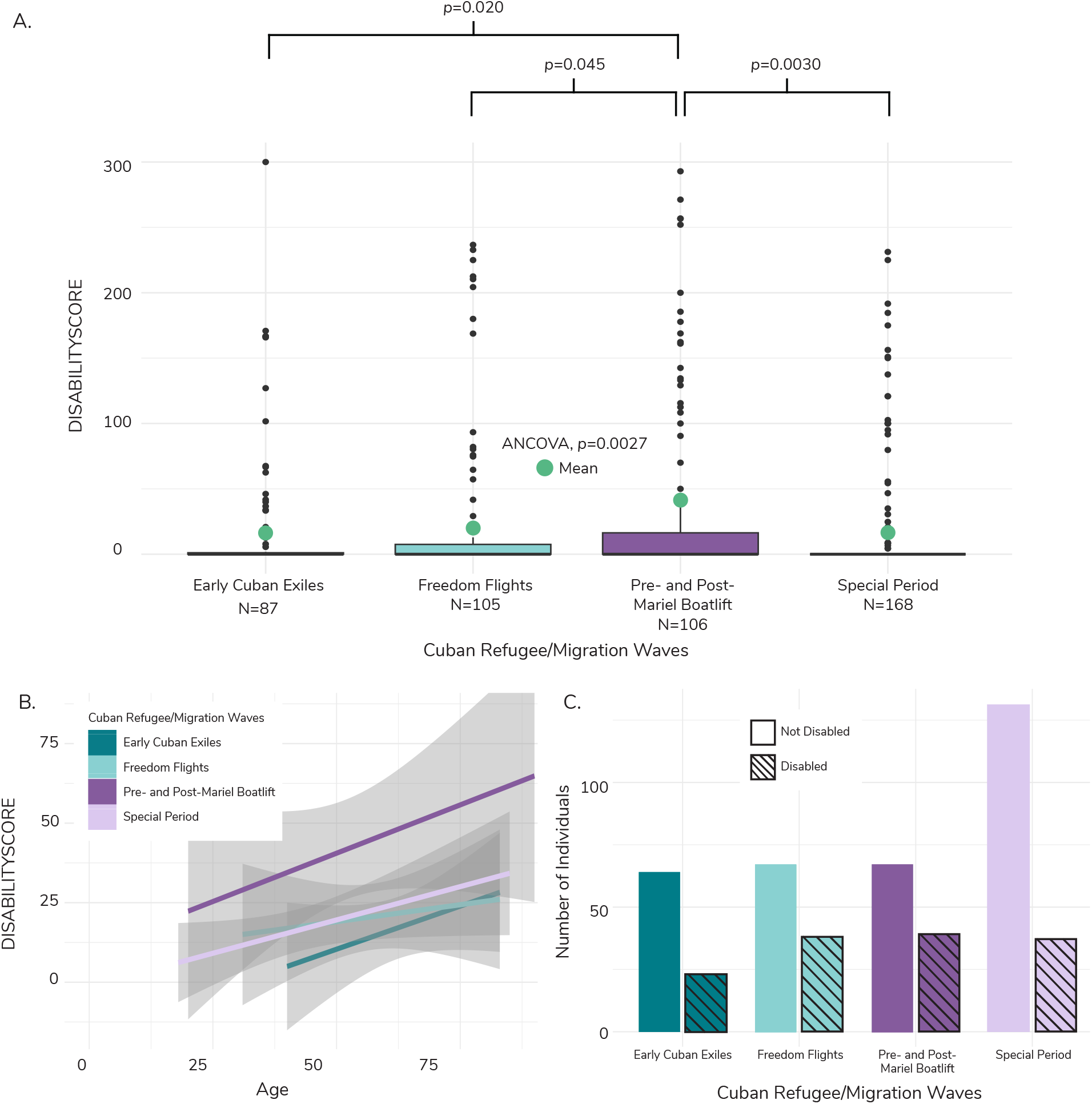
Composite disability score boxplot, trendlines, and frequency across Cuban refugee/migration wave. A) Boxplot of distribution of values and average for the composite disability score (DISABILITYSCORE) across Cuban refugee/migration waves. The green dots represent the weighted average, calculated using NLAAS sampling weights. We ran an ANCOVA to determine if there was a significant difference between means of different refugee/migration waves, while controlling for age, sex, and region. Refugee/migration wave was statistically signfiicant (p=0.0027). We ran a Tukey’s Honestly Signfiicant Difference (HSD) test to determine which averages were significantly different between waves. Average DISABILITYSCORE was significantly different between the Pre-and Post-Mariel Boatlift and all other refugee/migration waves, with p-values reported above the bracket for each respective refugee/wave comparison. B) Linear models with 95% confidence interval of the composite disability score (DISABILITYSCORE) plotted against age for each Cuban Refugee/Migration wave. The Pre- and Post-Mariel Boatlift model is offset higher relative to other waves, suggesting a higher level of disability across age. C) Barchart of individuals categorized as disabled within each Cuban refugee/migration wave. Individuals were categorized as disabled if they had a composite disability score (DISABILITYSCORE) greater than zero. Most individuals within each migration wave were categorized as not disabled. Disabled individuals are marked by the slanted lines for each wave.

We observed a similar pattern of increased instance of disability for Pre- and Post-Mariel Boatlift individuals when considering disability presence/absence rather than score. 36% of Early Cuban Exiles (N=23) were categorized as disabled, along with 57% of Freedom Flights (N=38), 58% of Pre- and Post-Mariel Boatlift (N=39), and 28% of Special Period individuals (N=37) (Figure 2C, Supplementary Figure 2). Being part of the Pre- and Post-Mariel Boatlift wave significantly predicted disability presence across cognition scores (logistic regression; *p*=0.036), mobility scores (*p*=0.040), role functioning scores (*p*=0.029), and sum disability score (*p*=0.018) (Figure 3). Individuals in the Pre- and Post-Mariel Boatlift wave had 2.93x, 2.37x, 2.31x, and 2.31x the odds seen in individuals from the Early Cuban Exiles for disability in cognition, mobility, and role functioning domains, and for the sum disability score, respectively (Early Cuban Exiles were used as the reference group for the logistic regression) (Figure 3).

**Figure 3.**
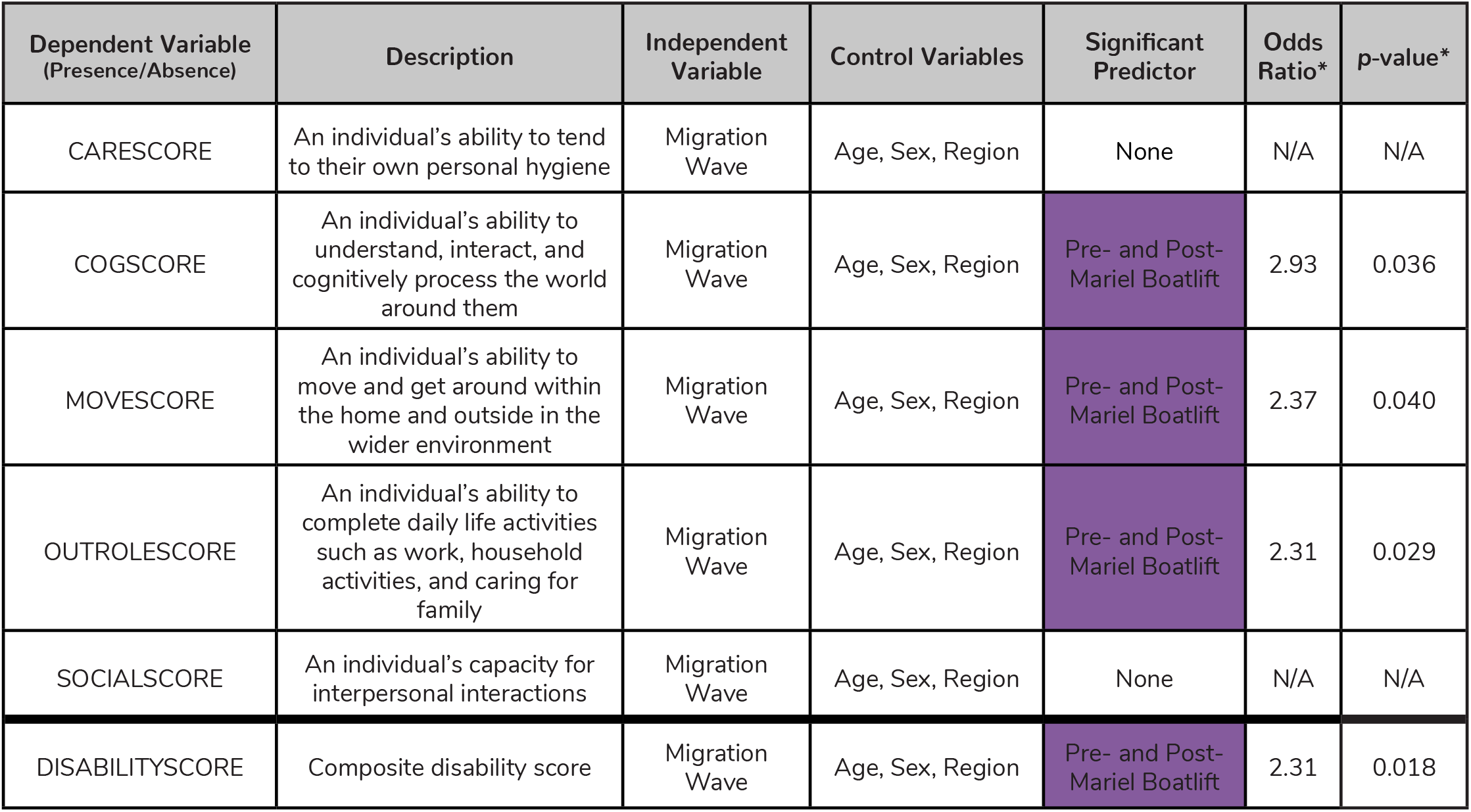
Logistic regression results for each health variable analyzed in this study. Here we show the results of the logistic regression models for each health measure converted to a binary variable. If a score across any domain was more than zero, those individuals were coded with the presence of a disability. We also include a general description of each health sure. We tested which migration wave may be associated with the presence of diability, while controlling for age, sex, and region across each model. All models include NLAAS sampling weights. *Odds ratios and p-values are reported for any predictor variable that was statistically significantly associated with the presence of disability (alpha < 0.05). *Odds ratios were generated relative to the odds of people having disabilities that fall within the Early Cuban Exiles migration wave. Summary statistics for each health measure are described further in Supplementary Figure 2.

### Pre- and Post-Mariel Boatlift individuals report experiencing relatively high discrimination

Experienced discrimination is known to be associated with adverse health outcomes (*25*). Among Cubans in the NLAAS, experienced Medical Discrimination (quantifying the amount of discrimination faced due to health problems within the last 30 days) significantly predicted the presence of disability within self-care scores (logistic regression; *p*=0.050), cognition scores (*p*=0.041), and social scores (*p*=0.023) (Supplementary Figure 3, Supplementary Table 1). Everyday Discrimination was marginally predictive of the sum disability score (*p*=0.091) and self-care scores (*p*=0.064), cognition scores (*p*=0.069), and social scores (*p*=0.073) (Supplementary Figure 3, Supplementary Table 1).

Medical discrimination was marginally significantly different across migration waves (ANCOVA, *p*=0.12; Figure 4); individuals from the Pre- and Post-Mariel Boatlift reported experiencing the highest average level of discrimination (1.49 ± 1.12) relative to the Early Cuban Exiles (1.00 ± 0.00), the Freedom Flights (1.34 ± 0.83), and the Special Period (1.14 ± 0.47). Experienced Everyday Discrimination (quantifying daily unfair treatment within the last year) averages did not differ significantly across waves (ANCOVA, *p*=0.84) (Figure 4, Supplementary Figure 3).

**Figure 4.**
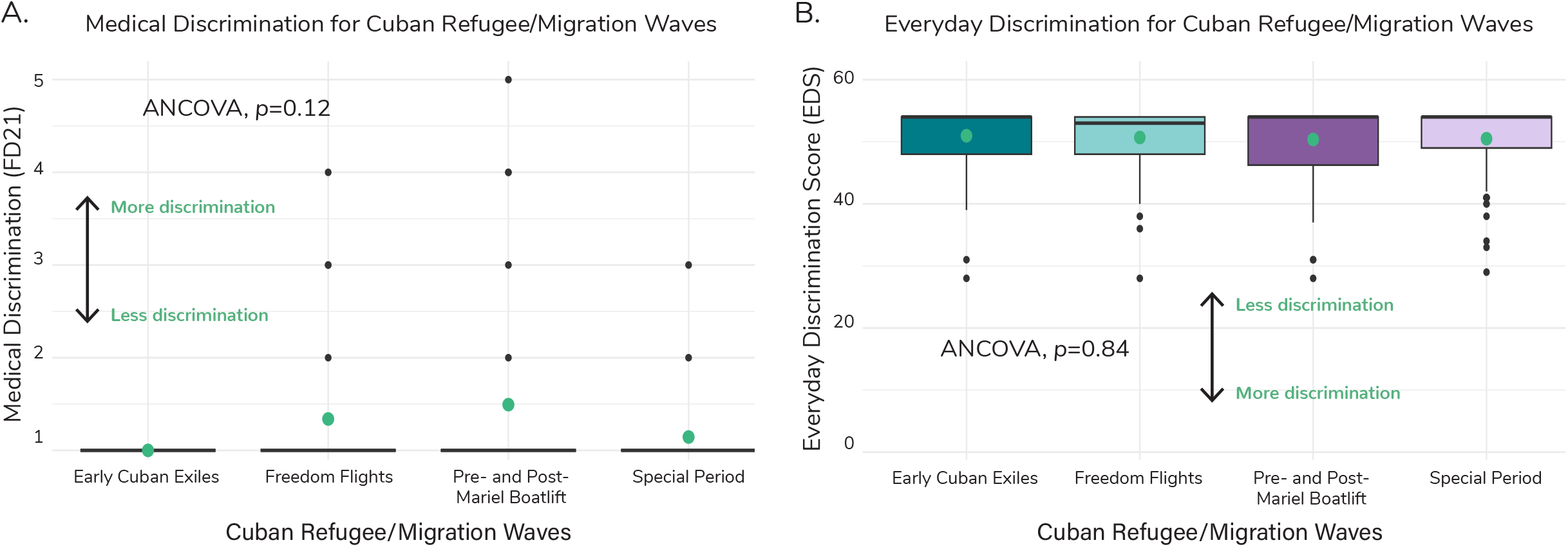
Medical Discrimination and Everyday Discrimination across Cuban Refugee/Migration waves. A) Boxplots of medical discrimination (coded as FD21 within NLAAS) by migration wave. Green dots represent the weighted average across each refugee/migration wave. We ran an ANCOVA to determine if there was a significant difference in means across the waves and found that there was no significant difference (*p*=0.12), as reported in panel A. For medical discrimination, a higher value represents more discrimination experienced and a lower value represents less. B) Boxplots of Everyday Discrimination Score (EDS) by migration wave. Green dots represent the weighted average across each refugee/migration wave. We ran an ANCOVA to determine if there was a significant difference in means across the waves and found that there was no significant difference (*p*=0.84), as reported in panel B. For EDS, a lower value represents more discrimination experienced and a higher value represents less.

### Mexican immigrants and Cuban immigrants show different patterns of disability among migration time frames

867 individuals in the NLAAS dataset at the time of our analysis were categorized as Mexican. Following our analyses with Cuban participant data, we removed individuals from the sample that were born in the United States (N=380) and those whose migration year was ambiguous for wave categorization (N=88). We categorized the remaining Mexican individuals based on the timeframe when they arrived in the United States, with n=21 individuals immigrating from 1953-1965 (corresponding to the Early Cuban Exiles), n=50 from 1965-1975 (corresponding to the Cuban Freedom Flights), n=115 from 1977-1987 (corresponding to the Cuban Pre- and Post-Mariel Boatlift), and n=197 from 1991-2002 (corresponding to the Cuban Special Period).

In stark contrast to the pattern observed among Cuban-Americans, for whom average disability was significantly greater among individuals who immigrated in the Pre- and Post-Marial Boatlift period (1977-1987) relative to earlier or later periods, sum disability scores for Mexican-Americans were instead highest for individuals immigrating to the U.S. from 1965-1975, corresponding to the Cuban Freedom Flights period (Figure 5), with sum disability scores (18.48 ± 40.21) 2.8x higher on average than individuals who migrated between 1977-1987 (6.42 ± 24.00; ANCOVA, *p*=0.006) and 11.5x higher on average than individuals that migrated between 1991-2002 (1.60 ± 6.65; *p*=0.00) (Supplementary Figure 4B).

**Figure 5.**
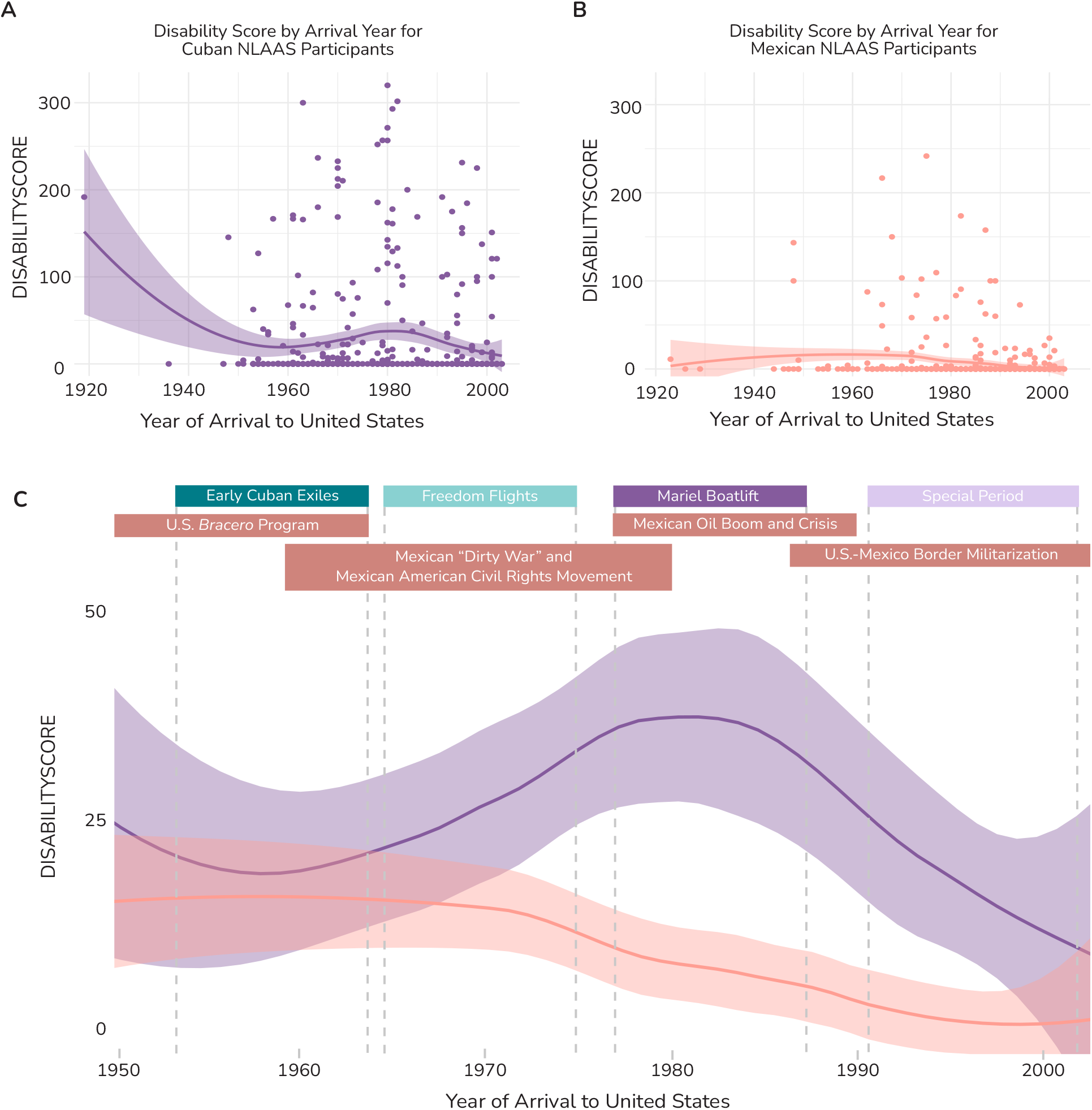
Composite disability scores per year of arrival to the United States for Cubans and Mexicans in NLAAS. (**A**) Scatter plot of DISABILITYSCORE for arrival year for Cubans within NLAAS. The purple line represents a LOESS regression, with the purple shaded region representing the 95% confidence interval. The regression peaks dramatically in 1920 (likely due to a single outlier that migrated in 1919) and again in 1981, around the time of Mariel Boatlift. (**B**) Scatter plots of average DISABILITYSCORE for arrival year for Mexicans within NLAAS. The orange line again represents a LOESS regression, with the orange shaded region representing the 95% confidence interval. The curve slightly peaks in 1972 and again around 1982. (**C**) Line plot of combined Cuban and Mexican LOESS regressions from A and B zoomed in from 1950 until 2003 and with a y-axis ranging (0, 1). Cuban migration waves and their associated time periods are marked at the top of the graph in their corresponding colors with vertical dashed lines. Events relevant to Mexican immigration and social history are also marked at the top of the graph in orange.

Medical discrimination was not significantly different across migration time periods for Mexican participants (ANCOVA, *p*=0.52). However, experienced Everyday Discrimination averages differed significantly across migration time periods (ANCOVA, *p*=0.0047), with individuals who migrated from Mexico to the U.S. from 1965-1975 having significantly higher scores (55.22 ± 4.18) relative to individuals that migrated between 1953-1964 (50.39 ± 7.87; ANCOVA, *p=*0.053), 1977-1987 (51.38 ± 8.57; ANCOVA, *p=*0.011), and 1991-2002 (50.98 ± 7.93; ANCOVA, *p*=0.0015) (Supplementary Figure 5).

## Discussion and Participant Interviews

The results from this study suggest that Pre- and Post-Mariel Boatlift wave immigrants have had higher overall instances *and* severities of disability relative to other Cuban refugee/migration waves, even when accounting for age and other factors. Individuals from this wave were also more likely to have experienced higher levels of some forms of discrimination. In contrast, a similar pattern of relatively elevated disability and experienced discrimination was *not* observed among Mexican immigrants to the U.S. over the same time period, suggesting that the results for Cuban immigrants may be culturally specific. Instead, Mexican participants who migrated to the U.S. between 1965-1975 reported higher discrimination scores and higher disability scores relative to other Mexican migration timeframes, including the 1977-1987 which corresponds to the Pre- and Post-Mariel Boatlift wave for Cuban immigrants to the U.S.).

As such, understanding specific socio-political backdrops in Cuba and the United States that may have impacted Cuban and Cuban American populations around the time of the Mariel Boatlift is thus essential to aid our understanding of how environmental variation may have been embodied by Cuban immigrants to contribute to the inter-wave health outcome disparities we observed. Interviews conducted as part of an ethnographic study aiming to investigate differences in ancestry, health, and narratives of immigration of individuals of Cuban descent in South Florida help us begin to consider the results of our NLAAS analyses in this context. Illustrative quotes from participants in this study are included in the discussion below.

### Social crises in Cuba as push factors for immigration during the late 1970s and early 1980s

In the late 1970s, Cuba experienced a severe economic crash (*26*). During this time, there was civil unrest, and Cubans rushed various embassies in Cuba to seek asylum. One of these rushes, known as the Peruvian Havana Embassy Crisis of 1980, resulted in over 10,000 Cubans attempting to seek asylum in Peru.

In response to this crisis, on April 20, 1980, the Cuban government opened the port of Mariel and told the Cuban people that anyone that wished to leave may do so. Over the next six months, over 125,000 Cubans fled to the United States, predominantly through boats and ships. In addition to anyone who expressed interest in leaving (who were considered traitors), the Cuban government expressed that anyone they deemed as “scum” - including imprisoned and queer people, sex workers, and those in mental institutions-should also leave. Discrimination faced by these marginalized groups in Cuba, then potentially exacerbated by adverse social conditions and experiences in the United States, may have contributed to the adverse health outcomes we observed in our study.

As one of the marginalized groups ostracized from the island, queer Cubans were a prevalent group migrating during the Mariel Boatlift. One participant who had many queer friends leave Cuba around the time of the Mariel Boatlift remarked:

“People who were gay in Cuba were marginalized. They would arrest them. They would arrest them. They abused them. That was terrible. They were sent to encampments. They did things to them that gay people don’t do…They couldn’t, as they say, come out of the closet. But there are gays where you can tell. And they, they beat them, they abused them. So, since they were marginalized in Cuba, that was one of the reasons why you could leave Cuba. Or because you were in prison, for stealing, for killing, for assault, what can I tell you? Or because you were gay.”

Another participant shared their father’s experience with an official while trying to board a boat during the Mariel Boatlift:

“I think [the official] told something…he probably like insulted [my dad]…and…[my dad] didn’t like that. [My dad] said something back and he just let the dog go and the dog, like, hurt my dad and attacked my dad as he was getting on the boat. And that was like…one of the last memories he has, like, vivid memories he has of leaving Cuba, was that moment.”

### Heightened racial discrimination and anti-immigrant sentiment in 1980 United States

Over the course of post-revolutionary Cuba-United States relations, Cubans have been allowed certain privileges in immigrating to the United States relative to other groups. For example, Cuban refugees could be admitted to a legal pathway to obtain U.S. residency and eventual citizenship that has not been readily available to other large, Latiné immigrant groups in the United States, such as refugees from Central America and Haiti (*14*). Yet, despite the existence of privileged legal avenues for any Cuban to become a member of United States society, social sentiment towards Cubans has not been consistent over the timespan of the four refugee waves explored in this paper.

First, racial hierarchies within the Cuban community have affected the Cuban refugee experiences within the United States. Ethnic and racial categories are mutable. The definition of what constitutes a particular race or ethnic group in society changes based on what country you are exploring these questions in, how individuals of that country identify one another in relation to these groups, and over time. Cubans embody these changes over the course of their post-revolutionary migration to the United States.

As discussed in other literature, nonwhite Cuban refugees from the Mariel Boatlift were found to have less economic opportunities and more difficulties establishing themselves in the Miami Cuban enclave than their white counterparts, and all members of this wave suffered similarly relative to individuals from earlier Cuban migration waves (*23*). The Mariel Boatlift and the Special Periods were both more racially and ethnically diverse (*27*), with our work indicating that they experienced higher burdens of discrimination relative to their earlier counterparts, possibly in part as a result.

One participant from the study recounted her father’s experiences of racial discrimination in the United States after coming during the Mariel Boatlift:

“My dad is the only person that like I constantly saw it, through his imprisonment. If he had been able to speak English and if he had been white presenting, I think his experience would have been wildly different. Because I’ve seen it in so many other people, I have friends who’ve had their white dads that speak English go to prison for kidnapping and get out within less than a decade. My dad is a non-violent crime and served life for it. It doesn’t add up. What is the missing link? Is it that he couldn’t navigate for himself?…They did their years, everybody who was in that car did less than five years. My dad did 25 to life for it. He was the only dark-skinned person in that car.”

Not only were Cubans of the Mariel Boatlift experiencing heightened discrimination, they were also facing stigmas associated with societal views of imprisoned people, sex workers, those in mental institutions, and queer people. During the year of the Boatlift, Miami was experiencing the first year of the AIDS epidemic and civil unrest as it related to the murdering of Arthur McDuffie, a Black insurance salesman who was killed by officers from the Dade County Public Safety Department.

Experienced prejudice and discrimination (*28*) and leaving one’s country due to social and/or economic strain can manifest in adverse overall health (*29, 30*), perhaps especially in combination. Although we cannot yet definitively conclude that the negative experiences disproportionately faced by Mariel Boatlift period refugees had direct, deleterious effects on their health, these factors seem likely to explain the relatively elevated levels of disability observed among these Cuban Americans relative to others, at least in part.

### Differential social experiences for Mexicans in the United States

Our study found that patterns of disability severity vary across different ethnic backgrounds. We examined disability severity within Mexican participants of the NLAAS and found that temporal variation in these scores was distinct from that observed among Cuban participants. Individuals who came to the U.S. from Mexico between 1965-1975 reported higher disability scores and experienced more discrimination relative to individuals from other timeframes, including the 1977-1987 timeframe corresponding to the Cuban immigration wave for which these scores were significantly higher than others. This result may reflect differences in the histories of immigrants’ countries of origin and/or divergent social experiences in their common host country.

Similar to Cuban migration, Mexican migration to the United States has been punctuated by social, political, and legislative events both in Mexico and the United States that served as variably driving or deterring forces for migrants (Figure 5C) (*31*). We considered factors that could potentially explain the higher disability and discrimination scores from Mexican participants that came to the U.S. in the 1965-1975 timeframe.

The United States Emergency Farm Labor Program, also known as the *Bracero* Program, began in 1942 with the intention of bringing Mexican laborers to the United States to supplement labor shortages as a result of the World War 2 (*30*). This program resulted in the signing of over 4.5 million work contracts that allowed Mexican migrants into the United States, mostly young men (*28*). In many cases, these men worked in dire conditions associated with low wages, dangerous working conditions, and inhospitable living conditions (*28*). The *Bracero* Program formally ended in 1964 (*32*).

In 1965, the United States passed the Immigration and Nationality Act, which sought to expand immigration to the United States from non-Western countries throughout the world. However, this act established strict limits on the number of migrants that could come from different countries/regions, imposing a cap of 120,000 total visas per year for countries within the Western Hemisphere, further reducing these visas to 20,000 per nation by 1976 (*28*). Contemporaneously, the Mexican “Dirty War”, lasting between the 1960s and 1980s, involved a single-party government controlled by the Institutional Revolutionary Party and resulted in violent repression of dissent within Mexico.

Within the United States, Mexican and Mexican American social movements rose prominently from the 1960s to the 1980s in response to adverse labor conditions, discriminatory policy changes, and substandard educational resources for people of Mexican descent in southern California (*28*).

Towards the end of the 1965-1975 timeframe highlighted by our analysis, a Mexican oil boom from the mid-1970s to early 1980s resulted in a reduced number of Mexican migrants entering the United States, though this boom was followed by a post-boom economic crisis (and thus increase in migration) lasting through the 1990s (*13, 14*). Towards the end of this economic crisis, the United States began an intense militarization of the U.S.-Mexico border to curtail migrants from entering the country (*33, 34*), which continues through the present day.

With respect to our results, we note that the Immigration and Nationality Act of 1965 resulted in a dramatic upshift of undocumented Mexican migration into the United States, thus creating a vulnerable population that did not previously exist in this capacity in the United States (*33*). Then, an oil-fueled economic shift in Mexico beginning in the mid-1970s altered this dynamic. The intervening window is demarcated by factors specific to Mexican immigrants, similar to the different specific factors that may have affected Cubans who immigrated to the U.S. in the pre- and post-Marial boatlift time period, potentially contributing to the stark differences we observed for participants from these two respective countries of origin.

### Heterogeneity exists both among and within Latiné groups

The first author is Cuban American, born to parents who migrated during different Cuban refugee/migration waves. Being able to zoom in on variation in lived experiences within Cuban refugees, and tying that social variation to health, is an example of how we can better appreciate our diversity and the real effects of this heterogeneity.

Oftentimes, Latiné people are grouped together in studies as a monolith, thus collapsing any potential measurable variation that we, as people that are descendants from these groups, know and appreciate. Our results robustly reject the monolithic treatment of Latiné peoples, at least when it comes to immigrant health outcomes. Specifically, we observed significant temporal variation in disability scores and experienced discrimination among Cuban immigrants, with relatively more disability and experienced discrimination during the years surrounding and including Mariel Boatlift of 1980. In turn, Mexican NLAAS participants had their own, but completely distinct from Cubans, temporal patterns of disability and experienced discrimination, further exemplifying this point.

Demonstrating that Latiné people can vary significantly both among countries of origin and even from within single countries of origin can hopefully inspire future studies that investigate heterogeneity within other similarly falsely homogenized ethnic and racial populations in the United States. As we show, this variation may have significant effects on overall health and well-being. By understanding the specific differential life experience circumstances in which adverse health outcomes arise (*35, 36*), we can better work towards collective solutions to mitigate these stressors for both current refugees/immigrants, and those who come to the United States in the future.

## Materials and Methods

### The National Latino and Asian American Study (NLAAS)

The National Latino and Asian American Study (NLAAS) is one of three studies under the Consortium in Psychiatric Epidemiology Studies (CPES) administered by the University of Michigan and funded by the National Institute of Mental Health (NIMH). NLAAS is a population-based survey sampling Latiné and Asian American individuals across the United States, including Alaska and Hawai’i (*35*). The purpose of the NLAAS was to collect information on the mental health needs and services used by individuals of Latiné and Asian descent, with a variety of additional data collected, including on socioeconomic variables, experiences of discrimination, and on physical health (*36*). This survey sampled 27,026 households with 4,469 completed responses. Respondents were selected based on a four-stage national area probability sample, with a special focus on individuals of Puerto Rican, Mexican, Cuban, Chinese, Filipino, and Vietnamese descent (*37*). The data for NLAAS are deposited in the Inter-university Consortium for Political and Social Research (ICPSR) online database under restricted access, where scientists interested in using the data can apply to use it via the ICPSR website (https://icpsr.umich.edu/) (*37*). We received permission from ICPSR for this project under Application #36467 titled “Variability in health outcomes among Cuban immigrants and Cuban Americans within the National Latino and Asian American Study (NLAAS)” on January 12, 2023.

As our analysis of this dataset uses de-identified data, our NLAAS analyses were deemed “not human subjects research” by the Penn State Office of Research Protections (STUDY00020329) and thus did not undergo further Institutional Review Board (IRB) review. We used the latest version of data available from the NLAAS at the time of analysis, which was last updated on March 23, 2016.

### Cuban participants within NLAAS

There are a total of N=577 individuals of Cuban descent present within the NLAAS dataset. For our research questions, we were specifically interested in understanding health among individuals that were first generation immigrants that came during different Cuban refugee/migration waves. We removed N=86 individuals from the sample that were born in the United States. The year individuals arrived in the United States was not a variable NLAAS collected. However, we were able to calculate year of migration based on three criteria: 1) year of birth, 2) age of arrival, and 3) year of participation in the study. There is a margin of error of ± one year due to the inclusion of year of birth. Using these migration years, we then categorized individuals into these waves using two methods: 1) the documented years historically associated with the cause of migration and 2) natural ebbs and flows within the data of year of migration. Based on these criteria, we created four bins of migration waves: 1) Early Cuban Exiles (1953-1964), Freedom Flights (1965-1974), Pre- and Post-Mariel Boatlift (1977-1986), and the Special Period (1991-2002). There is an additional wave, Operation Peter Pan (also known as *Operación Pedro Pan*), that was composed of over 14,000 Cuban children who were sent to the United States without a parent or guardian (*38*). In this analysis, we do not separate children that arrived during the Peter Pan flights into their own distinct wave, as they span from 1960 to 1962 and could not be distinguished from the Early Cuban Exiles migration wave in our dataset. There were some individuals who migrated during years that did not fall within the bins listed above; they were subsequently removed from the sample (N=25). A total of N=466 individuals were included in downstream analyses.

There were 87 Early Cuban Exiles, (mean age 62.49 ± 12.53), 105 Freedom Flights refugees (mean age 57.22 ± 13.88), 106 Pre- and Post-Mariel Boatlift refugees (mean age 51.72 ± 16.60) and 168 Special Period refugees (mean age 42.45 ± 15.60) (Figure 1, Supplementary Figure 1). There were 247 females and 219 males within the sample. NLAAS quantifies location in the United States via a code called Region, with four possible options: Midwest, Northeast, South, and West. Most individuals in the sample (N=436) were from the South. Full sample characteristics are reported in Supplementary Figure 1.

### Data analysis

Data characterization, statistical analyses, and data visualization were performed using the statistical software R and RStudio using the following packages: tidyverse, ggplot2, ggpubr, plyr, dplyr, patchwork, survey, mitools, lme4, car, and moments. Code for all analyses performed is available via the GitHub repository https://github.com/maggiehern/CubansNLAAS.

We noted that NLAAS collected a variable labeled SEX providing two options, male and female. We also noted that there was no formal collection of gender within the study. We hope future epidemiological studies include our wider understanding of sexual and gender diversity within their data collection methods to better represent any population of interest. This would include options for individuals to self-identify sex and the inclusion of intersex as a category. Additionally, social experiences (*39*) and health outcomes (*40*) can also differ by gender and gender expression and would also be critical to include in future studies.

### Health measures and statistics

We used several overall health measures present in the study to investigate health outcomes across Cuban migration waves. We specifically focused on measures of disability, as the consequences of adverse health may manifest in differences in an individual’s ability to function and participate in society (*41*). The disability measures used in this study include five of the six scores of the World Health Organization’s Disability Assessment Schedule II (WHODAS) (*42*). WHODAS scores were generated using a 36-item instrument that measures the level of impairment of an individual within the last 30 days of taking the assessment. Level of impairment was measured across five domains and given a distinct score for each. These domains are: self-care (CARESCORE), cognition (COGSCORE), mobility (MOVESCORE), social interactions (SOCIALSCORE), and life activities (OUTROLESCORE). The sixth domain, participation in community activities, was not collected as part of NLAAS. To generate these scores, participants rated their difficulty in completing tasks or participating in their social environment. For example, one of the questions related to mobility scores asks, “In the past 30 days, how much difficulty did you have in: Standing for long periods such as 30 minutes?” with the potential answers (and numerical values): none (0), mild (1), moderate (2), severe (3), and extreme (4). The numerical values were added together and converted to range from 0 to 100 with zero denoting no disability and 100 denoting full disability within each domain. These conversions can be calculated using the calculation matrix available on the WHO Disability Assessment Schedule website (*43*). WHODAS scores have been demonstrated to correlate with other validated health measures and can serve as proxy for physical and mental health disorders (*44*).

Self-care score measures an individual’s ability to tend to their own personal hygiene. Cognition score measures an individual’s ability to understand, interact, and cognitively process the world around them. Mobility score measures an individual’s ability to move and get around within the home and outside in the wider environment. Social interactions score measures an individual’s capacity for interpersonal interactions. Life activities score measures an individual’s ability to complete daily life activities such as work, household activities, and caring for family. Additionally, we created a composite disability score (denoted DISABILITYSCORE) by adding all WHODAS scores together for participants of Cuban descent in our sample to gain a full understanding of an individual’s level of disability. This overall measure of disability has been validated (*42*) and employed by other studies (*45*). This score had a possible range from 0-500. We checked whether DISABILITYSCORE was significantly correlated, and thus a good representation, of the various health measures that were used to create it by running correlation tests. We found that DISABILITYSCORE was significantly correlated with the CARESCORE (p < 2.2e-16, r = 0.63), COGSCORE (p < 2.2e-16, r = 0.62), MOVESCORE (p < 2.2e-16, r = 0.81), OUTROLESCORE (p < 2.2e-16, r = 0.91), and SOCIALSCORE (p < 2.2e-16, r = 0.58) (Supplementary Table 2). Additionally, we ran correlation tests to determine if the different domains of the WHODAS scores were correlated with one another (Supplementary Table 2). We found that all domains were correlated with one another, varying in their degree of explanatory power from r=0.32 to r=0.59 (Supplementary Table 2). Although these variables correlate, based on their method of collection, they measure different aspects of a person’s ability to engage in their everyday life (as shown through the WHODAS questionnaire). Additionally, Individuals that already have a disability in one domain may be more likely to have a disability in another (comorbidities). We calculated the mean and standard deviation for all health scores within each Cuban migration wave to gain insight into overall health.

We performed ANCOVAs for health measures across Cuban migration waves while controlling for age, sex, and region, as has been accounted for in other studies that have used NLAAS data (*46*). For ANCOVAs with *p*<0.05, we then used Tukey’s Honest Significant Difference (HSD) test for each pair of waves to determine if there was a significant difference in mean health scores.

Due to the large number of zero scores across WHODAS health scores, we dichotomized the variables by converting scores greater than zero to “present” for a disability and scores equal to zero as “absent”. This methodology has been validated by the WHO (*43*) and has been used in other studies that also use NLAAS data, due to the large frequency of zeroes present within many variables (not just health scores) in the dataset (*41, 47*). Using these dichotomized variables, we performed logistic regressions on all health measures using Cuban migration wave as the predictor variable and health score as the outcome variable while controlling for age, sex (reference category: male), and region (reference category: Northeast). We set the reference group as the Early Cuban Exiles for all logistic regression analyses. We report p-values and odds ratios for each logistic regression. All analyses incorporate sampling weights reported for NLAAS (variable name NLAASWGT). Including sampling weights in downstream analyses within epidemiological studies allows for our results to be made generalizable to populations of interest living in the United States.

### Discrimination indices and statistics

To gain a better understanding of some of the social factors that may be impacting overall health, we analyzed data in NLAAS relevant to perceived medical discrimination and perceived everyday discrimination. Medical discrimination (FD21) was measured in the NLAAS survey using a single question asking the participant to recall the amount of discrimination they faced due to health problems within the last 30 days with possible answers being none (coded as 1), a little (2), some (3), a lot (4) and extreme (5). Additionally, we explored perceived everyday discrimination using a nine-item survey within NLAAS (variables DS1A-DS1I; Supplementary Table 3). Questions were intended to understand various aspects of everyday life affected by experiencing prejudice. For example, question DS1A asks: With what frequency are you treated with less courtesy than others? Each question had six potential responses: almost every day (coded as 1), at least once a week (2), a few times a month (3), a few times a year (4), less than once a year (5), and never (6). Scores for these nine questions were added together to create a composite Everyday Discrimination Score (EDS) that ranged from 9-54, with lower scores indicating more discrimination and higher scores indicating less discrimination (*48, 49*) (Supplementary Table 3). We performed ANCOVAs using both discrimination measures, with Cuban migration wave as the predictor variable. We also performed logistic regressions with the various health measures described above as the dependent variables and amount of discrimination as the independent variables, to estimate the extent to which discrimination predicts the presence of a disability. We report averages and standard deviations for discrimination scores in Supplementary Figure 3.

### Mexican participants within NLAAS

There are a total of N=867 individuals of Mexican descent present within the NLAAS dataset. Mexican participants were categorized by on the time frames previously delineated by Cuban migration waves: 1) 1953-1964 (Early Cuban Exiles), 1965-1974 (Freedom Flights), 1977-1986 (Pre- and Post-Mariel Boatlift), and 1991-2002 (Special Period). There were 21 individuals that came between 1953-1964, (mean age 50.65 ± 10.31), 50 individuals that came between 1964-1974 (mean age 49.47 ± 10.01), 106 individuals that came between 1977-1987 (mean age 36.80 ± 8.62) and 168 individuals that came between 1991-2002 (mean age 28.58 ± 7.52). There were some individuals who migrated during years that did not fall within the bins listed above; they were subsequently removed from the sample (N=484). A total of N=383 individuals were included in downstream analyses. There were 199 females and 184 males within the sample. Most individuals in the sample (N=217) were from the West.

We performed ANCOVAs for health measures and experienced discrimination scores across migration time frames while controlling for age, sex, and region, as done with the Cuban participants. For ANCOVAs with *p*<0.05, we then used Tukey’s Honest Significant Difference (HSD) test for each pair of waves to determine if there was a significant difference in mean health scores.

### Mexican and Cuban disability by age per year of migration

Our original approach for this project involved delineating Cuban participants into migration timeframes that made sense relative to distinct sociopolitical events or conditions within Cuba and the United States. However, we wanted to understand if disability scores varied in another pattern outside of the patterns we predicted. To visualize this, we plotted disability scores divided by age for all individuals that migrated to United States from Cuba across all migration years present in the NLAAS dataset. We did the same for Mexicans NLAAS participants. We also plotted a LOESS (locally-estimated scatterplot smoothing) regression for each participant group.

### Ethnographic data: The Cuban Immigration and Health Study (CIHS)

We use ethnographic data from the Cuban Immigration and Health Study (CIHS) to contextualize the results from thus research study. In the CIHS, we investigated the diversity that exists within individuals of Cuban descent in terms of their ancestry, lived experiences, and health outcomes. We used a snowball sampling methodology of individuals that live in the South Florida area, were of Cuban descent (either they, their parents, or their grandparents immigrated from Cuba after the Communist Revolution) and were 18 years of age or older. Ethnographic data for this project was collected via two approaches: surveys and semi-structured interviews. These tools were used to collect general demographic information (including age, family characteristics, income and other measures relevant to understanding socioeconomic position), year of migration, self-identified ethnicity and self-identified race, perceived ethnic discrimination, access to healthcare, medical history, adverse health risk factors, and narratives of arrival to the United States. Qualitative data from surveys was used to understand the general health profile of individuals from different migration waves in context with the results of the semi-structured interviews. Interviews were conducted in either English or Spanish, depending on the preference of the participant. Spanish language interviews were translated by the first author. During the semi-structured interview, we asked participants to recall frequency and instances of ethnic and racial discrimination, stories on hardships (or lack of) faced when arriving in the United States, social support and sentiments they received and perceived when arriving, and descriptions of overall health and wellbeing. These interviews seek to identify potential causal mechanisms for adverse health outcomes among Cuban migrants in general or among different Cuban migration waves, adding much needed context to studies like NLAAS. This project has been approved by Penn State’s Institutional Review Board (IRB) under study number STUDY00016932.

## Supporting information

All Supplementary Figures and Tables

## Data Availability

All data used for this study were provided by the Inter-university Consortium for Political and Social Research and are available online at https://icpsr.umich.edu

https://github.com/maggiehern/CubansNLAAS

## Acknowledgments

The authors would like to thank the participants of the National Latino and Asian American Study and the Cuban Immigration and Health Study, whose time and effort made this project possible. The authors would also like to thank Dr. Mary Shenk, Dr. Lindsay Fernández-Rhodes, and Dr. Mark Shriver for their analytical advice and assistance in editing this manuscript. The authors also thank Emily Davenport for suggesting the comparative Mexican immigrant analysis and Cynthia Gonzalez, Rocio Cisneros, and Esther Muñoz for their helpful conversations about Mexican and Mexican American history. This research was supported by funding from the National Science Foundation Graduate Research Fellowship Program grant DGE1255832 (to M.H.) and the National Institutes of Health grant 1F99HG012711-01 (to M.H.). Any opinions, findings, and conclusions or recommendations expressed are those of the authors and do not necessarily reflect the views of the funding agencies listed above.

