## Supplementary material for "Latiné immigrant heterogeneity: Striking health differences among Cuban refugee/migration waves to the United States": All Supplementary Figures and Tables

| Variables | Early Cuban Exiles | Freedom Flights | Pre- and Post-Mariel Boatlift | Special Period |
|---|---|---|---|---|
| N | 87 | 105 | 106 | 168 |
| Age | 62.49 ± 12.53 | 57.22 ± 13.88 | 51.72 ± 16.60 | 42.45 ± 15.60 |
| Females (N) | 48 | 65 | 51 | 83 |
| Males (N) | 39 | 40 | 55 | 85 |
| Northeast (N) | 3 | 14 | 6 | 5 |
| South (N) | 83 | 90 | 100 | 163 |
| West (N) | 1 | 1 | 0 | 0 |

**Supplementary Figure 1. Descriptive statistics for Cuban subsample of NLAAS.** Here, we report general descriptive statistics for the Cuban subsample of the NLAAS study. Average age and standard deviation was generated while incoroporating sampling weights from the NLAAS dataset (see Methods). Note: The individuals included in these analyses are those that met the cutoffs for the migration waves of interest for this study, therefore not all individuals that identify as Cuban in NLAAS are present in these statistics.

| Health Measures | Early Cuban Exiles | Freedom Flights | Pre- and Post-Mariel Boatlift | Special Period |
|---|---|---|---|---|
| N | 87 | 105 | 106 | 168 |
| CARESCORE | 1.85 ± 10.08 | 1.07 ± 6.43 | 3.47 ± 12.68 | 0.82 ± 5.76 |
| COGSCORE | 1.08 ± 5.86 | 3.17 ± 12.10 | 4.67 ± 13.47 | 1.71 ± 8.58 |
| MOVESCORE | 5.61 ± 17.63 | 4.60 ± 14.73 | 8.52 ± 20.51 | 3.63 ± 14.26 |
| OUTROLESCORE | 7.96 ± 22.54 | 10.35 ± 26.88 | 21.74 ± 39.42 | 9.45 ± 25.42 |
| SOCIALSCORE | 0.64 ± 3.39 | 1.39 ± 7.05 | 3.18 ± 12.61 | 0.81 ± 5.14 |
| DISABILITYSCORE | 17.14 ± 45.83 | 20.58 ± 53.38 | 41.59 ± 81.76 | 16.41 ± 44.13 |
| Disabled (N) | 23 | 38 | 39 | 37 |
| Not Disabled (N) | 64 | 67 | 67 | 131 |

**Supplementary Figure 2. Health variable summary statistics for Cuban subsample of NLAAS.** This table reports the average, standard deviation, and N (where applicable) for the health measures of interest in this study for the Cuban cohort. These results were generated while incoroporating sampling weights from the NLAAS dataset (see Methods). Note: The individuals included in these analyses are those that met the cutoffs for the migration waves of interest for this study, therefore not all individuals that identify as Cuban in NLAAS are present in these statistics. The composite disability score (DISABILITYSCORE) was calculated by adding together all five WHODAS scores for each individual. Individuals were categorized as disabled if they had a composite disability score greater than zero.

| Health Measures | Early Cuban Exiles | Freedom Flights | Pre- and Post-Mariel Boatlift | Special Period |
|---|---|---|---|---|
| Medical Discrimination | 1.00 ± 0.00 | 1.34 ± 0.83 | 1.49 ± 1.12 | 1.14 ± 0.47 |
| Everyday Discrimination | 50.98 ± 5.18 | 50.69 ± 4.71 | 50.37 ± 5.74 | 50.50 ± 5.05 |

**Supplementary Figure 3. Discrimination scores summary statistics for the Cuban subsample of NLAAS.** This table reports the averages and standard deviations for Medical Discrimination and Everyday Discrimination scores of the Cuban cohort. Averages and standard deviations were generated while incorporating sampling weights from the NLAAS dataset (see Methods). Note: The individuals included in these analyses are those that met the cutoffs for the migration waves of interest for this study, therefore not all individuals that identify as Cuban in NLAAS are present in these statistics.

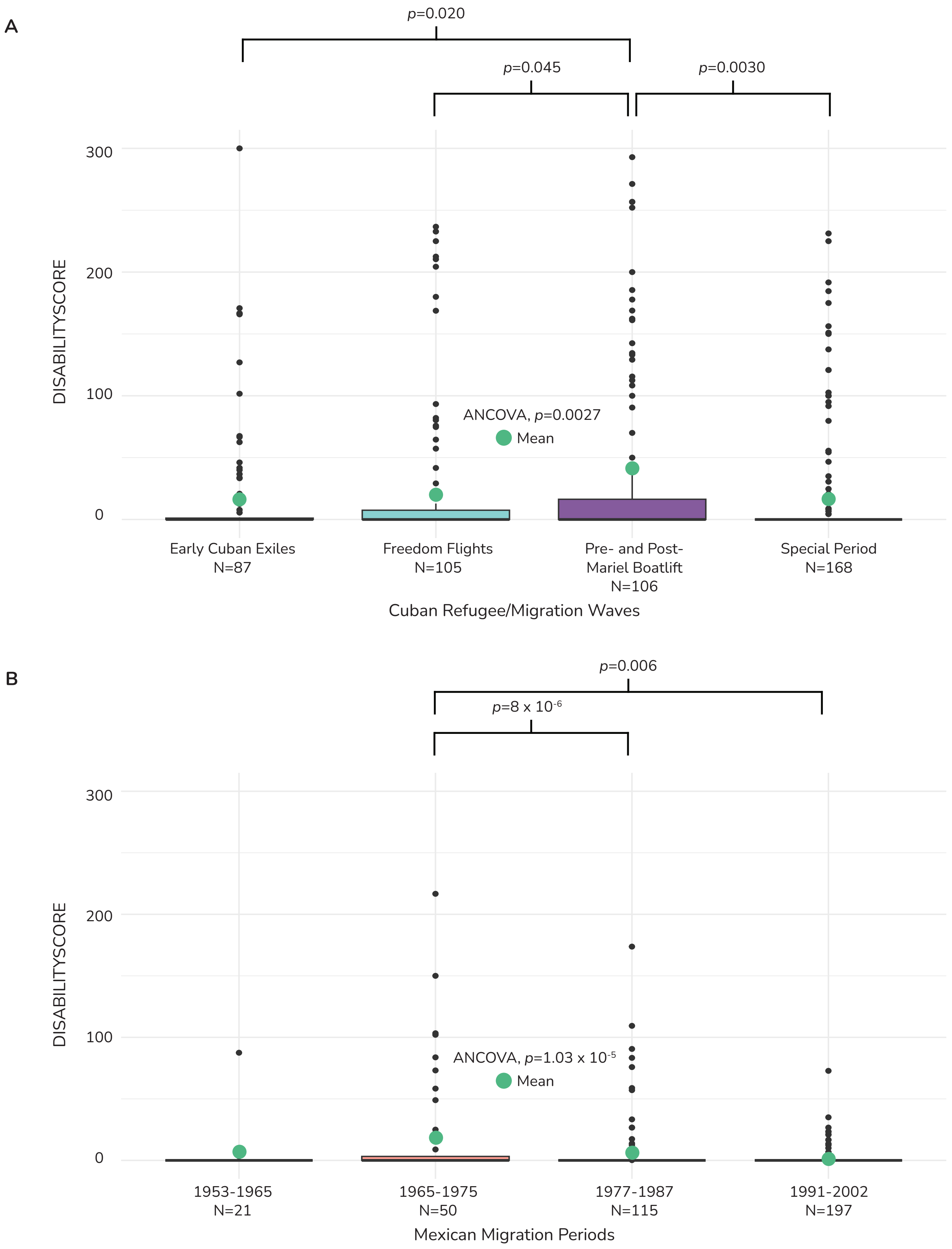

**Supplementary Figure 4. Composite disability scores for Cubans and Mexicans in NLAAS.** (**A**) Boxplot of distribution of values and average for the composite disability score (DISABILITYSCORE) across Cuban refugee/migration waves. (**B**) Boxplot of distribution of values and average for the composite disability score (DISABILITYSCORE) across Mexican migration periods that correspond to Cuban refugee/migration waves.

| Health Measures | 1953-1964 | 1965-1975 | 1977-1987 | 1991-2002 |
|---|---|---|---|---|
| Medical Discrimination | 0.00 ± 0.00 | 1.63 ± 1.15 | 1.17 ± 0.59 | 1.49 ± 0.58 |
| Everyday Discrimination | 50.39 ± 7.87 | 55.22 ± 4.18 | 51.38 ± 8.57 | 50.98 ± 7.93 |

**Supplementary Figure 5. Discrimination scores summary statistics for the Mexican subsample of NLAAS.** This table reports the averages and standard deviations for Medical Discrimination and Everyday Discrimination scores of the Mexican cohort. Averages and standard deviations were generated while incorporating sampling weights from the NLAAS dataset (see Methods).

| **Health Score** | CARESCORE | COGSCORE | MOVESCORE | OUTROLE-SCORE | SOCIALSCORE | DISABILITY-SCORE |
|---|---|---|---|---|---|---|
| CARESCORE | | | | | | |
| COGSCORE | $r = 0.32$<br>$p < 2.2e-16$ | | | | | |
| MOVESCORE | $r = 0.56$<br>$p < 2.2e-16$ | $r = 0.39$<br>$p < 2.2e-16$ | | | | |
| OUTROLESCORE | $r = 0.40$<br>$p < 2.2e-16$ | $r = 0.45$<br>$p < 2.2e-16$ | $r = 0.59$<br>$p < 2.2e-16$ | | | |
| SOCIALSCORE | $r = 0.43$<br>$p < 2.2e-16$ | $r = 0.54$<br>$p < 2.2e-16$ | $r = 0.38$<br>$p < 2.2e-16$ | $r = 0.37$<br>$p < 2.2e-16$ | | |
| DISABILITYSCORE | $r = 0.63$<br>$p < 2.2e-16$ | $r = 0.62$<br>$p < 2.2e-16$ | $r = 0.81$<br>$p < 2.2e-16$ | $r = 0.91$<br>$p < 2.2e-16$ | $r = 0.58$<br>$p < 2.2e-16$ | |

**Supplementary Table 1. Correlation coefficients for NLAAS health measures analyzed in this study.** Here, we report correlation coefficents and p-values for each Pearson's Correlation Test conducted between health measures used within this study. All health measures were significantly positively correlated with one another.

| Question | Variable | Range | Scale |
|---|---|---|---|
| Frequency treated with less courtesy than others | DS1A | 1-6 | 1: Almost everyday<br>2: At least once a week<br>3: A few times a month<br>4: A few times a year<br>5: Less than once a year<br>6: Never |
| Frequency treated with less respect than others | DS1B | | |
| Frequency received poorer restaurant service than others | DS1C | | |
| Frequency people act like you are not smart | DS1D | | |
| Frequency people act afraid of you | DS1E | | |
| Frequency people act like you are dishonest | DS1F | | |
| Frequency people act better than you | DS1G | | |
| Frequency called names/insulted | DS1H | | |
| Frequency threatened/harrassed | DS1I | | |

**Supplementary Table 2. Everyday Discrimination Score (EDS) survey questions.** Survey questions used to generate the Everyday Discrimination Score (EDS) used as a measure of overall discrimination as part of this study. Participants were the frequency with which they experience discrimination on an everyday basis, with six potential responses: almost everyday, at least once a week, a few times a month, a few times a year, less than once a year, or never. These responses were used to create a numerical value, with a lower value indicating more discrimination and a higher value indicating less discrimination. The scores for survey items DS1A-DS1I were added together to a create a compostion Everyday Discrimination Score, with a range of 9-54.

| Dependent Variable (Presence/Absence) | Description | Independent Variable | Control Variables | Odds Ratio | p-value |
|---|---|---|---|---|---|
| CARESCORE | An individual's ability to tend to their own personal hygiene | Medical Discrimination (FD21) | Age, Sex, Region | 1.89 | 0.050 |
| COGSCORE | An individual's ability to understand, interact, and cognitively process the world around them | Medical Discrimination (FD21) | Age, Sex, Region | 1.85 | 0.041 |
| MOVESCORE | An individual's ability to move and get around within the home and outside in the wider environment | Medical Discrimination (FD21) | Age, Sex, Region | N/A | 0.85 |
| OUTROLESCORE | an individual's ability to complete daily life activities such as work, household activities, and caring for family | Medical Discrimination (FD21) | Age, Sex, Region | N/A | 0.33 |
| SOCIALSCORE | An individual's capacity for interpersonal inter-actions | Medical Discrimination (FD21) | Age, Sex, Region | 3.87 | 0.023 |
| DISABILITYSCORE | Composite disability score | Medical Discrimination (FD21) | Age, Sex, Region | N/A | 0.94 |
| CARESCORE | An individual's ability to tend to their own personal hygiene | Everyday Discrimination (EDS) | Age, Sex, Region | N/A | 0.064 |
| COGSCORE | An individual's ability to understand, interact, and cognitively process the world around them | Everyday Discrimination (EDS) | Age, Sex, Region | N/A | 0.069 |
| MOVESCORE | An individual's ability to move and get around within the home and outside in the wider environment | Everyday Discrimination (EDS) | Age, Sex, Region | N/A | 0.25 |
| OUTROLESCORE | An individual's ability to complete daily life activities such as work, household activities, and caring for family | Everyday Discrimination (EDS) | Age, Sex, Region | N/A | 0.33 |
| SOCIALSCORE | An individual's capacity for interpersonal inter-actions | Everyday Discrimination (EDS) | Age, Sex, Region | N/A | 0.073 |
| DISABILITYSCORE | Composite disability score | Everyday Discrimination (EDS) | Age, Sex, Region | N/A | 0.091 |

**Supplementary Table 3. Logistic regression results for discrimination scores.** Here we show the results of the logistic regression models for each health measure converted to a binary variable. If a score across any domain was more than zero, those individuals were coded with the presence of a disability. We test if medical discrimination and everyday discrimination scores are associated with the presence of diability, while controlling for age, sex, and region across each model. All models include NLAAS sampling weights.
